# Stitch with Which? Absorbable vs Nonabsorbable Suture for Skin Closure – A meta-analysis

**DOI:** 10.1101/2023.03.02.23286680

**Authors:** Dev Desai

**Author notes:** Contact no.

## Abstract

**Background:** It has been taught as a fundamental value that for skin sutures and wound closure, non-absorbable sutures should be used without many research papers supporting its higher efficacy with decreased would infection, decreased wound dehiscence and decreased skin scarring. The optimal suture material type still remains a myth.

**Objective:** This study aimed to compare the outcomes of absorbable versus non-absorbable sutures for skin closure

**Methods:** A Systemic-review was performed with randomized controlled trials (RCTs) and Cohort studies that compared outcomes of absorbable versus non-absorbable sutures for skin closure.

**Results:** A total of 5096 patients in 27 RCTs analyzed. There was no significant difference between absorbable sutures and non-absorbable sutures in the incidence of wound infections, scar formation and wound dehiscence. The results of both groups are quite comparable with nil heterogeneity.

**Conclusions:** Absorbable sutures for skin closure were not inferior to nonabsorbable sutures. It should be recommended due to its great cost and time savings. Well-designed RCTs with sufficient follow-ups are needed to adequately clarify whether better cosmetic results can be achieved using intradermal absorbable sutures.

## Introduction

For most surgeons, skin closure is crucial. Every year, thousands of surgeries are conducted, with almost every one including skin closure. Just a small improvement in skin closure can result in significant expense savings and time savings. Sutures, staples, and surgical glues are commonly used to close wounds on the skin. In comparison to suture closure, staples have a much increased risk of wound infection. (1) Nonabsorbable Suture Group (NAG) was considered as the standard method for skin closure. (2) (3) (4) However, it must be painfully removed after suturing, necessitating a second medical appointment and lost work days. In youngsters, suture removal might be challenging, especially if they have bad recollections of the initial repair. With minimal reactivity and great tensile strength, absorbable sutures are commonly utilized in elective surgery to enhance aesthetic results, save expenses, and increase patient satisfaction. (2) (5) hence, Absorbable Sutures Group(AG) challenged the nonabsorbable suture for the position of ideal skin closure suture.

Wound issues can lengthen your hospital stay or force you to return. According to specialists, absorbable sutures can also be utilized in contaminated incisions and are preferred in contaminated wounds. (5) (6) However, no significant differences between absorbable and nonabsorbable sutures with respect to wound infection were reported in multiple studies. (2) (4) (7). A study showed that with absorbable sutures, a higher rate of wound infection is seen (8), but papers about cosmetic outcome and scar formation were contradicting as some paper showed better outcomes (9) (10) in compared to non-absorbable whereas some showed cosmetic equivalence. (11) Also, better patient satisfaction and low cost was seen with absorbable sutures. (12) (13)

Rapid skin healing along with a satisfactory aesthetic appearance while limiting the risk of complications are the benchmarks of efficient wound closure. Many previous investigations on wound closure with absorbable or nonabsorbable sutures yielded mixed results. Only one meta-analysis, published in 2007 (14), examined absorbable and nonabsorbable sutures in traumatic lacerations and surgical wounds. However, it was based on only seven trials with small sample sizes (less than 356 patients in the absorbable sutures group), and five of the seven randomized controlled trials (RCTs) included in this meta-analysis were low-quality studies with a Jadad score of less than 3, which may not be suitable for meta-analysis. It came to the following conclusion: To compare the efficacy of absorbable vs nonabsorbable sutures for wound closure, large methodologically sound RCTs were required. Since then, more well-designed RCTs with a substantial number of additional patients have been conducted to assess the efficacy of absorbable suture. As a result, this study conducted a meta-analysis of RCTs to assess the effects of absorbable and nonabsorbable sutures for skin closure.

**(Chart 1).**
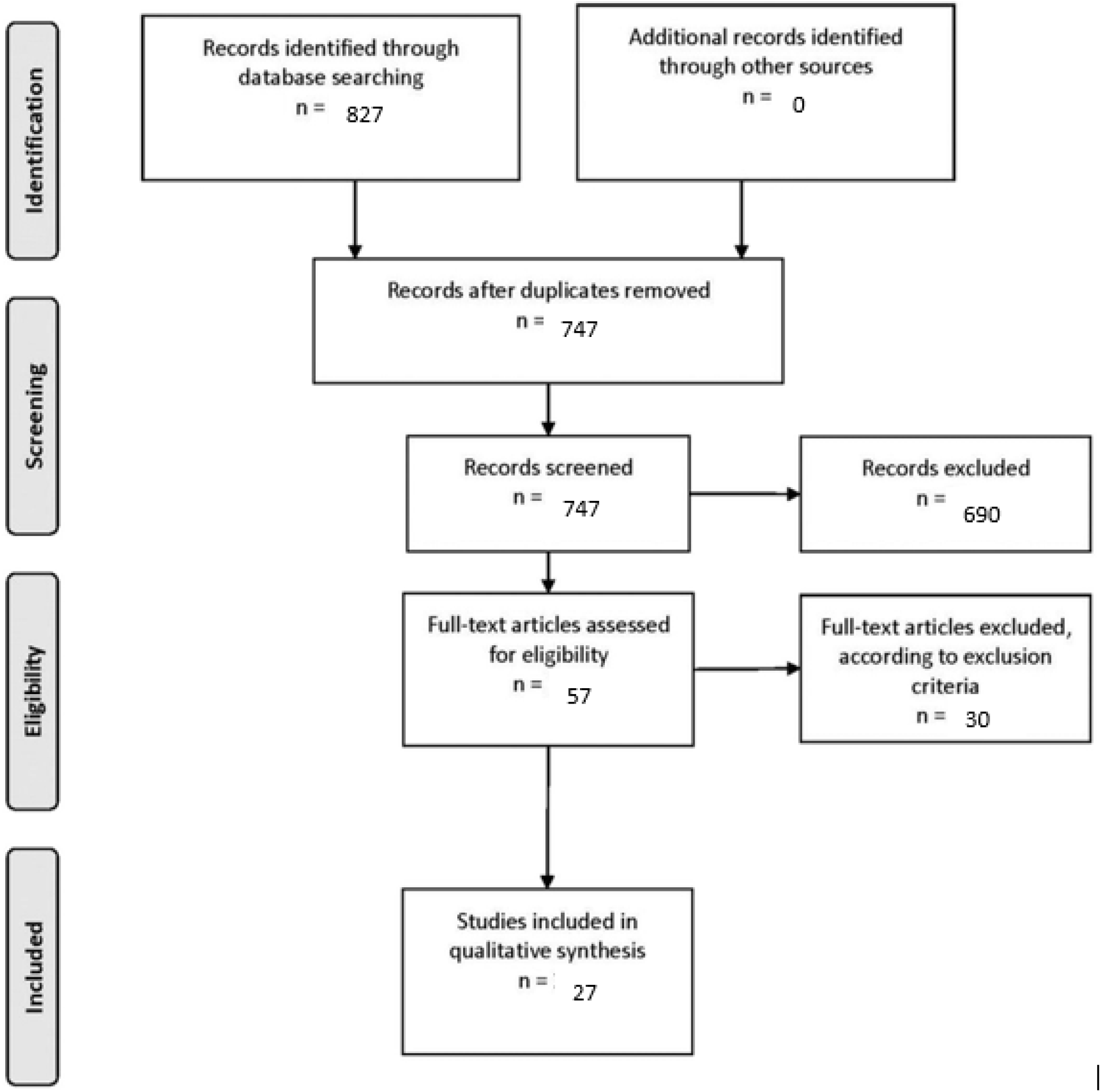
PRISMA Flow Chart

## Methodology

### Data source

A computerized search was undertaken in PubMed, EMBASE, OVID, Web of Science, the Cochrane Library, Google Scholar, and the Controlled Trials Meta Register. “Absorbable” and “nonabsorbable” were text keywords. Additional studies were found by manually searching the reference lists of pertinent retrieved publications. The only language allowed was English. Human investigations were the only ones that yielded results.

### Eligibility Criteria

The researchers looked for randomized controlled studies that compared absorbable vs nonabsorbable sutures in the treatment of skin closure. Along with Abstracts, letters, comments, editorials, expert opinions, reviews without original data, and case reports, Research papers were also excluded from the analysis if (1) it was impossible to extract the appropriate data from the published articles; (2) there was significant overlap between authors, institutes, or patients in the published literatures; (3) the measured outcomes were not clearly presented in the literatures; (4) the measured outcomes were not clearly presented in the literatures; and (5) articles were written in non-English.

### Study Identification

All titles and abstracts discovered by the search approach were reviewed by the author.

Relevant complete publications were collected for detailed evaluation; two non-author independent reviewers separately assessed them for eligibility criteria.

### Data extraction

Each qualified manuscript was reviewed by two independent reviewers separately. The number of patients, their age, gender, operational procedures, wound types, closure methods, and the incidence of wound infections, dehiscence, and scar formation were all collected from each manuscript. Conflicts were addressed by further discussion or consultation with the author and a third party. The modified Jadad score was used to measure the study’s quality. (15)

### Outcome Measures

The frequency of wound infection and cosmetic effects following surgery were our primary outcomes. The incidence of wound dehiscence was among the secondary outcomes studied.

### Statistical analysis

All of the information was gathered and placed into software for analysis. To analyze important clinical outcomes, fixed- or random-effects models were used to generate mean difference, standardized mean difference (SMD), odds ratios, and relative risk (RR) with 95 percent confidence intervals (CIs). Statistical heterogeneity was measured with the χ^2^; P < 0.100 was considered as a representation of significant difference. I^2^ greater than or equal to 50% indicated the presence of heterogeneity. (16) The fixed-effects model was used to aggregate the findings in the absence of statistically significant heterogeneity. According to DerSimonian and Laird, the random-effects model was utilized once heterogeneity was proven. (17) To compare the findings of surgical wound infection and aesthetic outcomes, subgroup analyses and/or one study deleted meta-analysis were used. Funnel plots were used to assess potential publication bias based on the prevalence of wound infection after surgery. A statistically significant difference was defined as P<0.05.

## Results

### Description of studies

827 citations were identified with initial search from which 27 eligible papers were identified (27 Randomized control trials). Some articles didn’t had full data accessible yet needed data could be retrieved from already published paper by Al-Abdullah et al. (14) The description of all articles is summarized in Table 1.

**Table 1:**
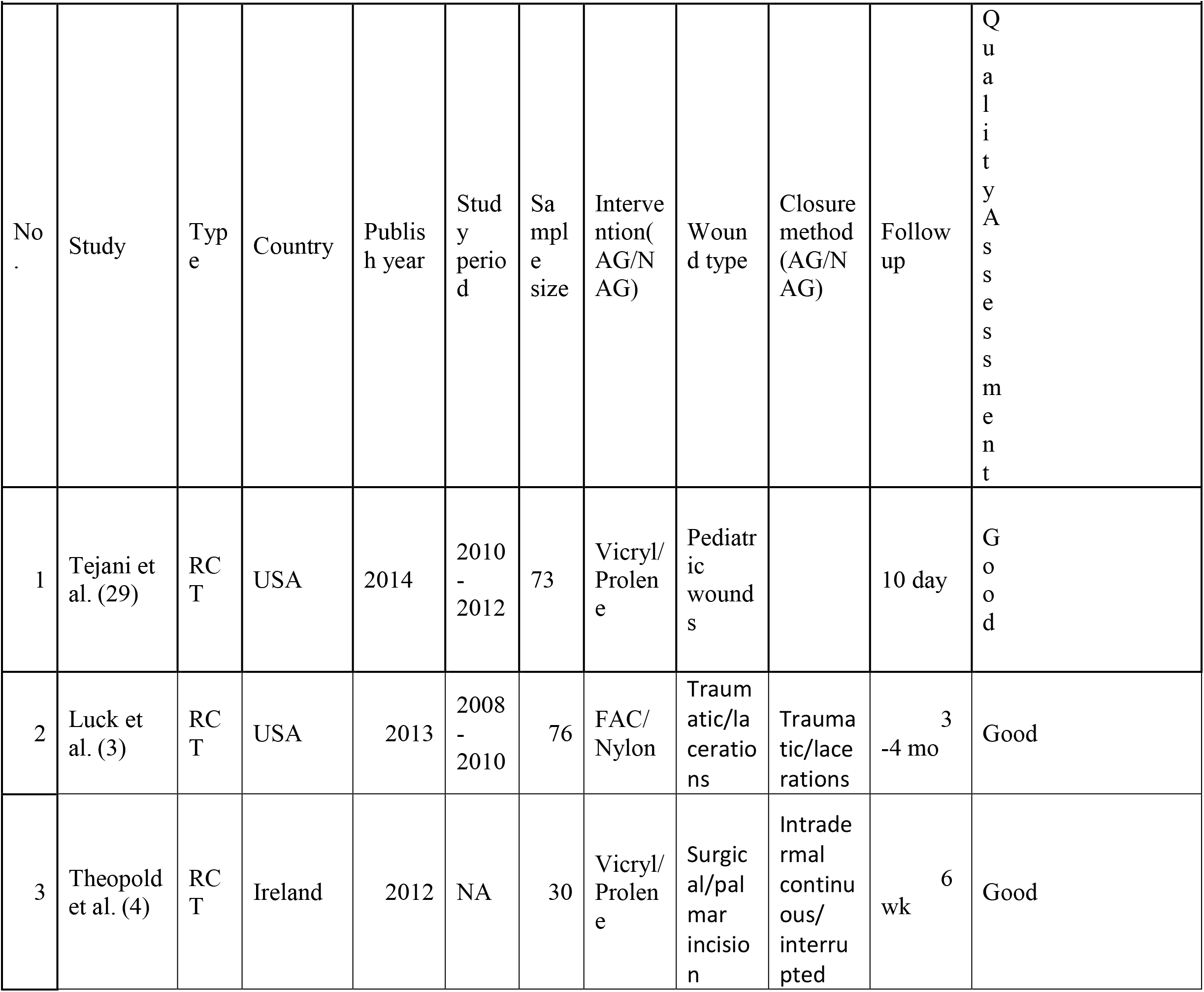
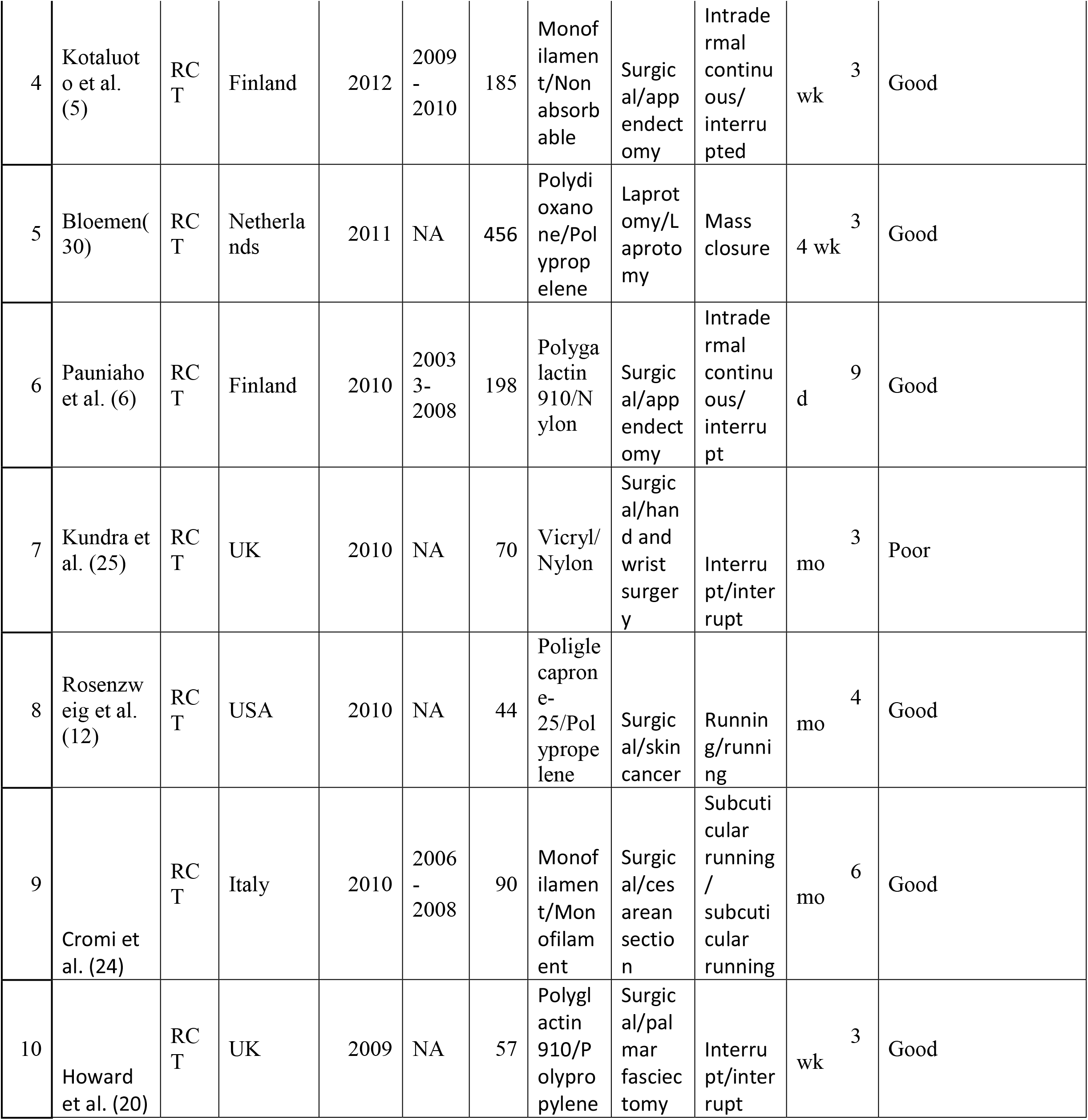
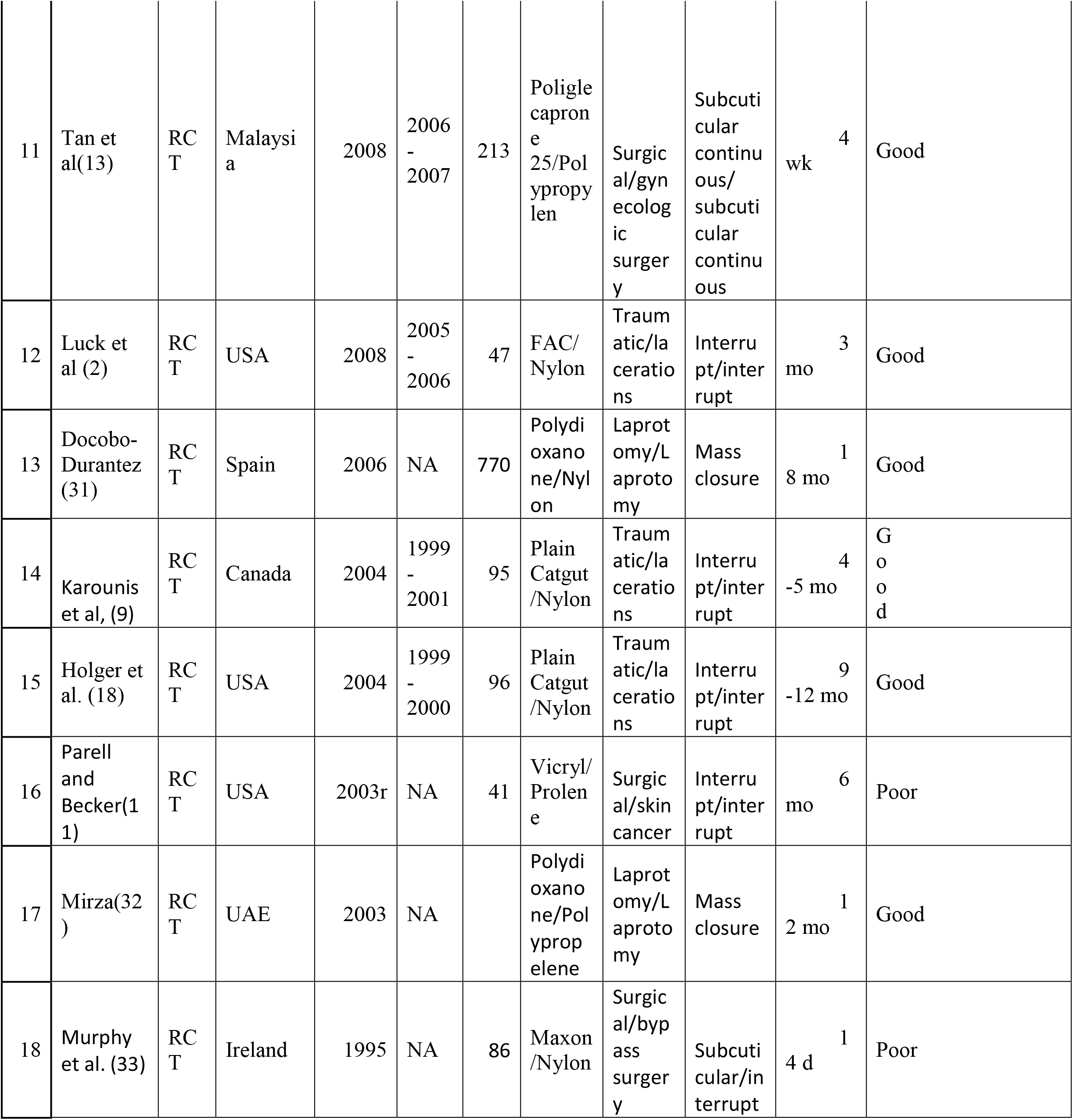
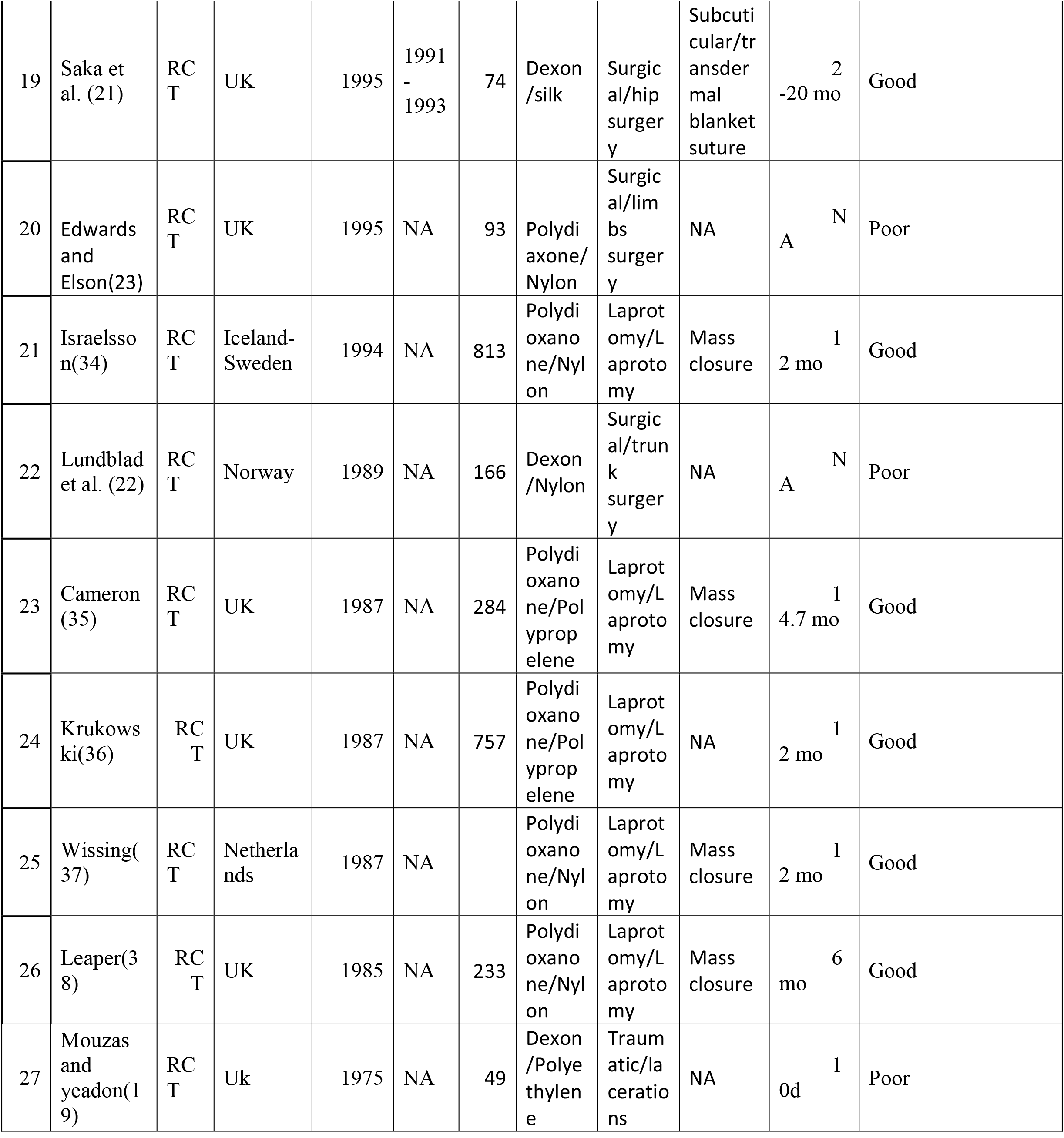
Description of studies

27 RCTs with total of 5096 patients either in AG (Absorbable group) or NAG (Non-Absorbable group). The baseline characteristic of both group was similar. Some information regarding age in some papers (11) (12) (14) (18) (19) (20) (21) were not sufficient along with in some papers information under gender distribution was not clear (22) (19, 23) (20) (21).

Methodological Quality has been summarized in Table 1. Researches have shown to be using Computer generated randomization (5, 13) (24) (20) (18) sealed envelope (2-4, 25) and coin toss (6) (12) for randomization. Quality was assessed and evaluated with the help of modified Jadad score (15) with a possible score from 0 to 7 (highest level of quality). Jadad score of 4 to 7 was defined as “Good” whereas Jadad score of 3 or less was defined as “Poor”. “Good” quality was determined in 13 studies and “poor” quality was determined in 6 studies.

### Wound Infection

Wound Infection after suturing was taken as a parameter to be considered by every research paper. No wound infection in AG were reported by only 2 out of all 28 research papers. (9, 20) and in NAG also only 2 research papers showed no wound infection. (18, 24). With an RR of 1.01 {95 percent CI, 0.83–1.23}), a meta-analysis of all included trials found no significant difference in the incidence of wound infections between absorbable and non-absorbable sutures. {χ^2^ = 25.23(P=0.56), Z= 0.09 (P=0.93), I^2^ = 0%}. Subgroup analysis was carried out in papers that had a high Jadad score. It was discovered that there was no significant difference between AG and NAG in terms of postoperative infection, independent of the patients’ age, wound type, or closure procedures. In the previously indicated analyses, no indication of heterogeneity between included studies was found.

**Comparison 1 (1.1):**
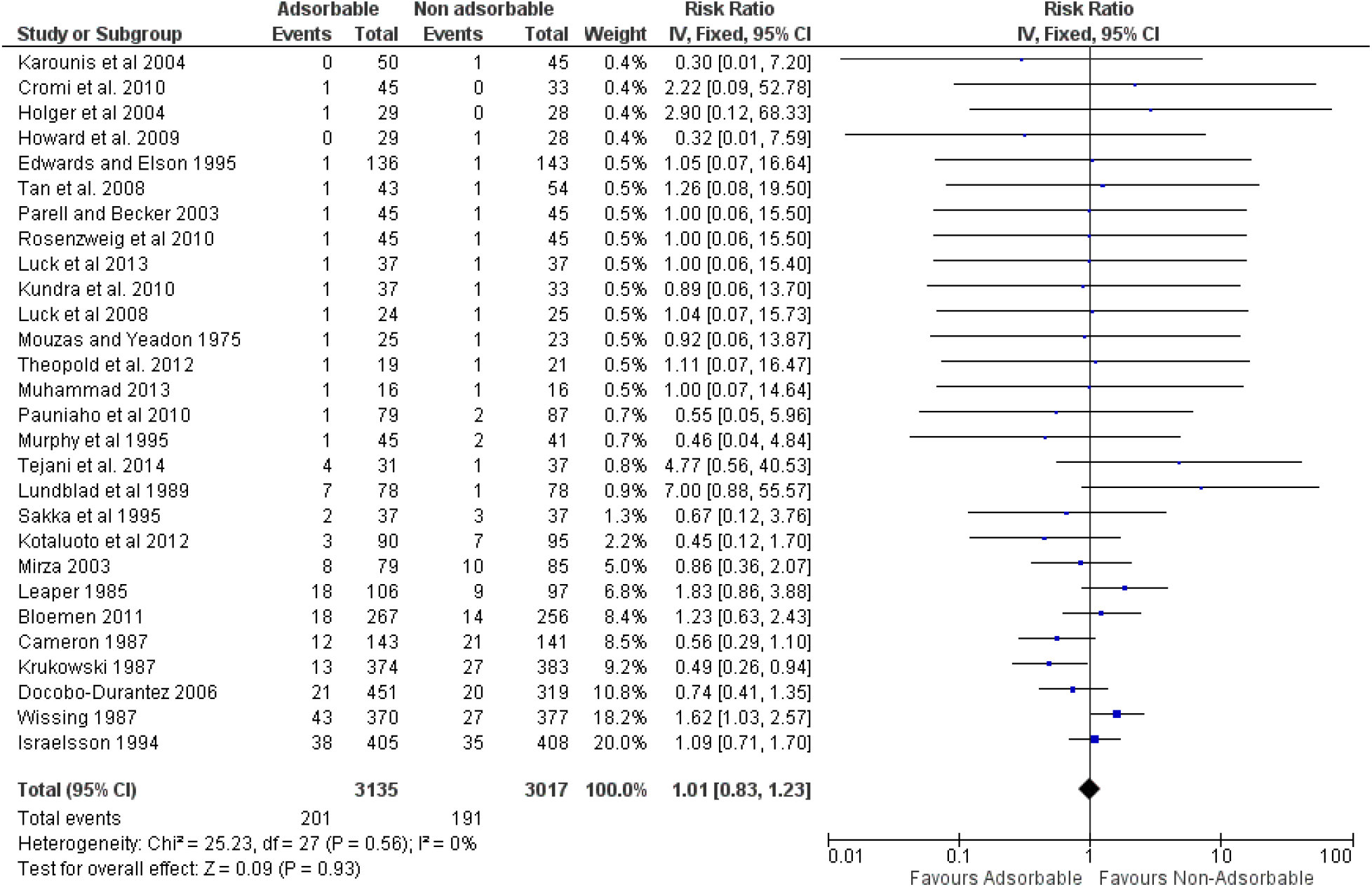
Wound infection (All papers) (Chart 2)

**Comparison 1(1.2):**
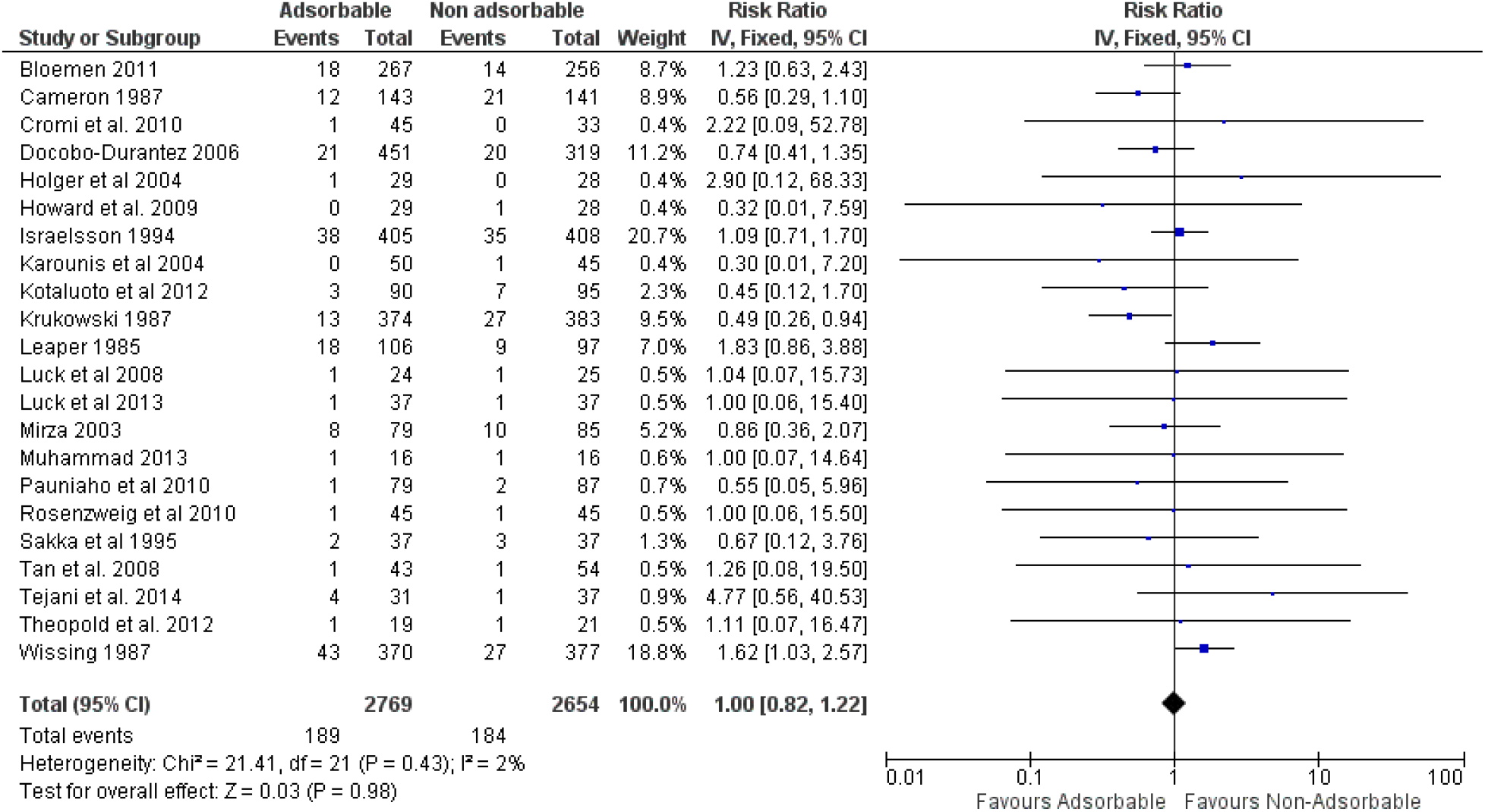
Wound infection (With high jaded score) (Chart 3)

### Wound Dehiscence

Many papers reviewed didn’t have sufficient information. Out of total 27 papers, 17 papers had data about wound dehiscence. There was no significant difference between absorbable and nonabsorbable sutures with an RR of 1.00 (95% CI, 0.66–1.52). No heterogeneity between studies was identified. {χ^2^ = 17.87(P=0.27), Z= 0.02 (P=0.99), I^2^ = 16%}

**Comparison 2:**
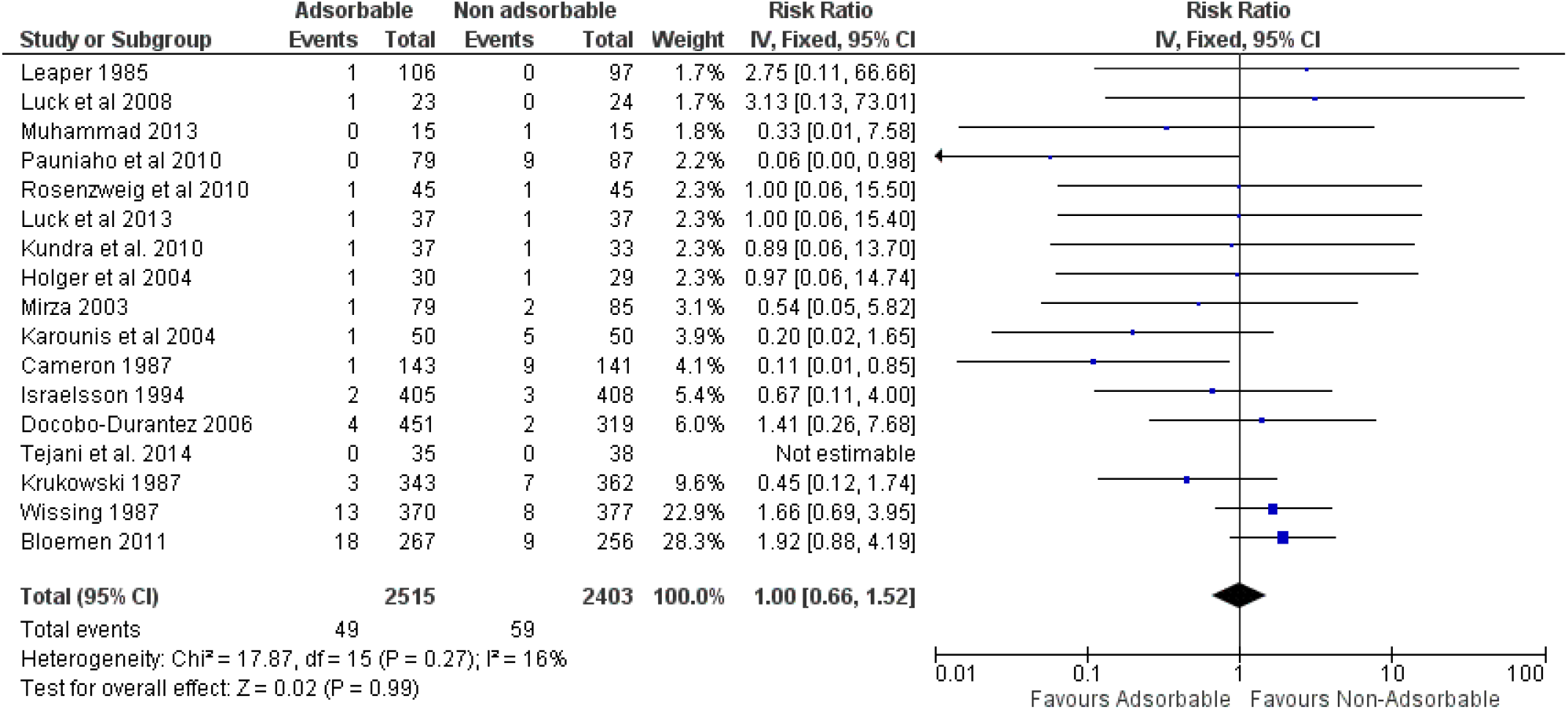
Wound Dehiscence (Chart 4)

### Skin Scar

Overall Cosmetic outcome was measured on 1 month to 12-month time spread in almost 9 studies. (2-4, 9, 12, 13, 18, 21, 24) and was assessed using Observer Scar Assessment Scale, Visual analog Scale (VAS), Vancouver Scar Scale and other cosmetic Assessment scales. There was no significant difference found between both groups one in AG and other in NAG. Statistical analysis show that RR was of 1.07 (95% CI, 0.46–2.46). No heterogeneity between studies was identified. {χ^2^ = 4.17(P=0.52), Z= 0.15 (P=0.88), I^2^ = 0%}

**Comparison 3:**
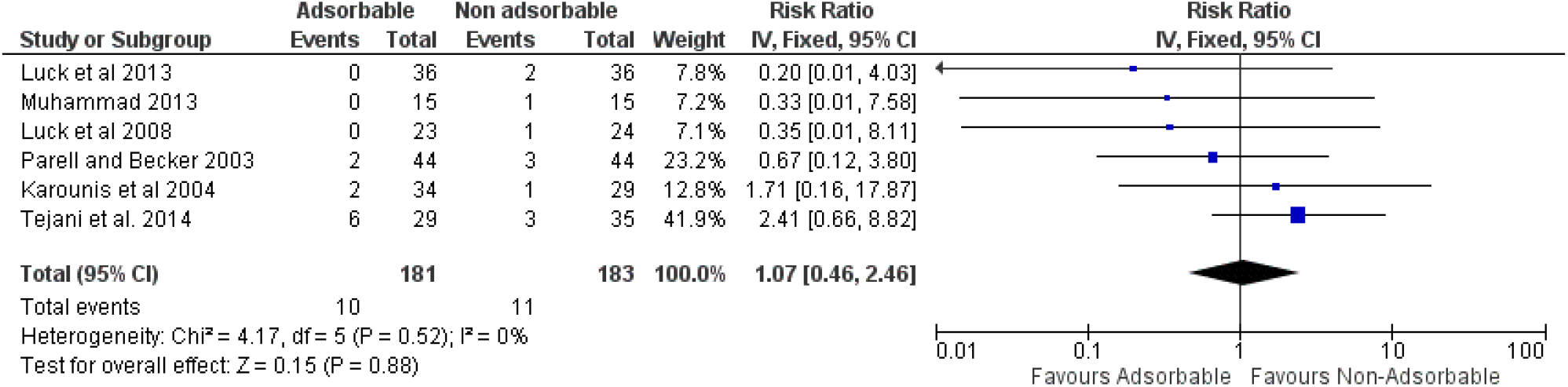
Skin Scar (Chart 5)

## Discussion

Skin closure has typically not been done using absorbable sutures. Several recent investigations, however, have shown that it may be used safely in skin closure. The debate over skin closure using absorbable sutures continues, and considerable effort need to be done to resolve it in an evidence-based manner. An absorbable suture for skin closure was found to be a viable alternative to typical non-absorbable sutures in our meta-analysis; absorbable sutures should be suggested.

Although cosmetics is an important aspect, other significant outcomes such as infection and wound dehiscence must be addressed when comparing suture materials for laceration healing. (14) A number of key criteria influence the method of skin closure chosen. Some studies have suggested that absorbable sutures may cause more frequent suture responses, compromising wound healing. (12) However, a research by Kotaluoto et al (5) found that AG had fewer wound problems than NAG. But in other trials, no significant changes in wound appearance or infection rates between absorbable and nonabsorbable sutures were found. No significant difference between AG and NAG was seen in our meta-analysis for skin closure. The result’s stability and reproducibility were further validated by a sensitivity analysis. Subgroup analysis was also done in RCTs with high Jadad scores. It also found no significant difference between AG and NAG in postoperative infections, independent of the patients’ age, wound type, or closure procedures. It also found no significant difference between AG and NAG in terms of post-suturing wound infections, independent of the patients’ age, wound type, or closure procedures. Wound dehiscence has to be taken into account when comparing suture materials for skin closure, however our investigation revealed no significant differences. As a result, in terms of postoperative morbidity, absorbable sutures for skin closure were not inferior to nonabsorbable sutures.

It’s unknown if absorbable suturing can produce improved aesthetic results. Cosmetic results and scar development were not substantially different between absorbable and nonabsorbable sutures, according to our meta-analysis. There might be some bias as skin scar condition and what to consider a skin scar with different scales like VAS is subjective. A better created methodology trying to differentiate between absorbable and non-absorbable suturing at intradermal level can show a story entirely different where absorbable suture might come as the best suturing material. It is subject to the amount of follow-ups, the level of skin suturing and the modality and condition while the sutures were taken. (13, 21, 24)

One of the biggest advantages of absorbable sutures versus nonabsorbable sutures is that they don’t need to be removed as often. In the treatment of juvenile facial lacerations, one economic analysis comparing the economic cost of absorbable sutures, nonabsorbable sutures, and tissue adhesives was published. (26) This study looked at the costs of equipment, pharmaceuticals, health-care worker time, and parental income loss for follow-up visits, assuming that ED overhead, registration fees, and parental time off work for the first ED visit were all identical. (14). Both patients and health care professionals benefitted from the use of Absorbable sutures for skin closure(5). Hargreaves (27) found that using absorbable sutures in everyday surgery might save a lot of money without sacrificing clinical efficacy or safety.

According to Phillips and Eilbert (28), using a single plain gut suture and laceration repair kit saved $19.54 over using a single nylon suture for the same treatment. Also, it eliminates the need of follow up visits and also the distress and pin and the negative social anxiety towards suture removal can be avoided especially in cases of children. Considering the advantages, Absorbable sutures should be advised. (14)

Some of the trials included in this meta-analysis had patients and assessors who were not properly blinded to the outcome method. In several experiments, allocation concealment was not specified explicitly. Heterogeneity between existing research, a lack of a single evaluation standard, and imperfect and incomplete recollection may all impair cosmetic ratings. To reduce variation in future clinical studies, standardized cosmetic assessment and definition for distinct clinical criteria are required. Furthermore, in principle, the varied closure types of deeper layers and skin should be random, similar, and adequately controlled in RCTs using absorbable and nonabsorbable sutures. However, this clinical difference exists objectively, and most studies did not indicate how to close the deeper layers, thereby introducing heterogeneity into our metaanalysis. To resolve the ambiguity that different types of shutting deeper layers may impact the aesthetic effect, well-designed RCTs are required.

### Conclusion

In conclusion, Non-absorbable sutures are no better than Absorbable sutures when it comes to preventing wound infection or wound dehiscence or the scar that they create. Given its better cost and time saving properties, despite common practice of using non-absorbable sutures, use of Absorbable sutures is recommended. With better patient compliance, it can lead to a healthier recovery. Whether to use absorbable sutures in classical patterns or to use it intradermal for approximation of skin wounds is still a question and more papers with ideal methodology is required to actually get a significant result.

## Data Availability

All data produced in the present study are available upon reasonable request to the authors

## References

1. Smith TO SD, Mann C,. Sutures versus staples for skin closure in orthopaedic surgery 2010 [Available from: https://pubmed.ncbi.nlm.nih.gov/20234041/.

2. Luck RP FR, Eyal D,. Cosmetic outcomes of absorbable versus nonabsorbable sutures in pediatric facial lacerations 2008 [Available from: https://pubmed.ncbi.nlm.nih.gov/18347489/.

3. Luck R TT, Gerard J,. Comparison of cosmetic outcomes of absorbable versus nonabsorbable sutures in pediatric facial lacerations 2013 [Available from: https://pubmed.ncbi.nlm.nih.gov/23714755/.

4. Theopold C PS, Dempsey M,. A randomised controlled trial of absorbable versus non-absorbable sutures for skin closure after open carpal tunnel release 2012 [Available from: https://pubmed.ncbi.nlm.nih.gov/21987279/.

5. Kotaluoto S PS, Helminen M,. Wound healing after open appendectomies in adult patients: a prospective, randomised trial comparing two methods of wound closure 2012 [Available from: https://pubmed.ncbi.nlm.nih.gov/22669400/.

6. Pauniaho SL L-VT, Helminen MT,. Non-absorbable interrupted versus absorbable continuous skin closure in pediatric appendectomies. 2010 [Available from: https://pubmed.ncbi.nlm.nih.gov/21044931/.

7. Fosko SW HD. Surgical pearl: an economical means of skin closure with absorbable suture. 1998 [Available from: https://pubmed.ncbi.nlm.nih.gov/9704837/.

8. Foster GE HE, Hardcastle JD,. Subcuticular suturing after appendicectomy. 1977 [Available from: https://pubmed.ncbi.nlm.nih.gov/68225/.

9. Karounis H GS, Eisman H,. A randomized, controlled trial comparing long-term cosmetic outcomes of traumatic pediatric lacerations repaired with absorbable plain gut versus nonabsorbable nylon sutures. 2004 [Available from: https://pubmed.ncbi.nlm.nih.gov/15231459/.

10. Pereira JL VG, de Albuquerque LA,. Skin closure in vascular neurosurgery: a prospective study on absorbable intradermal suture versus nonabsorbable suture 2012 [Available from: https://pubmed.ncbi.nlm.nih.gov/23050208/.

11. Parell GJ BG. Comparison of absorbable with nonabsorbable sutures in closure of facial skin wounds 2003 [Available from: https://pubmed.ncbi.nlm.nih.gov/14623686/.

12. Rosenzweig LB AM, Ho J,. Equal cosmetic outcomes with 5-0 poliglecaprone-25 versus 6-0 olypropylene for superficial closure 2010 [Available from: https://pubmed.ncbi.nlm.nih.gov/20653727/.

13. Tan PC MS, Omar SZ,. Absorbable versus nonabsorbable sutures for subcuticular skin closure of a transverse suprapubic incision 2008 [Available from: https://pubmed.ncbi.nlm.nih.gov/18639876/.

14. Al-Abdullah T PA, Fergusson D,. Absorbable versus nonabsorbable sutures in the management of traumatic lacerations and surgical wounds: a meta-analysis. 2007 [Available from: https://pubmed.ncbi.nlm.nih.gov/17505281/.

15. Banares R AA, Rincon D, et al. Endoscopic treatment versus endoscopic plus pharmacologic treatment for acute variceal bleeding: a meta-analysis. 2002 [Available from: https://pubmed.ncbi.nlm.nih.gov/11870374/.

16. Higgins JP TS, Deeks JJ,. Measuring inconsistency in metaanalyses 2003 [Available from: https://pubmed.ncbi.nlm.nih.gov/12958120/.

17. DerSimonian R LN. Meta-analysis in clinical trials 1986 [Available from: https://pubmed.ncbi.nlm.nih.gov/3802833/.

18. Holger JS WS, Hale DB,. Cosmetic outcomes of facial lacerations repaired with tissue-adhesive, absorbable, and nonabsorbable sutures 2004 [Available from: https://pubmed.ncbi.nlm.nih.gov/15258862/.

19. Mouzas GL YA. Does the choice of suture material affect the incidence of wound infection? A comparison of dexon (polyglycolic acid) sutures with other commonly used sutures in an accident and emergency department 1975 [Available from: https://pubmed.ncbi.nlm.nih.gov/1106807/.

20. Howard K SA, Morris A,. A prospective randomised trial of absorbable versus non-absorbable sutures for wound closure after fasciectomy for Dupuytren’s contracture. 2009 [Available from: https://pubmed.ncbi.nlm.nih.gov/19687084/.

21. Sakka SA GK, Abdulah A,. Skin closure in hip surgery: subcuticular versus transdermal. A prospective randomized study. 1995 [Available from: https://pubmed.ncbi.nlm.nih.gov/8571773/.

22. Lundblad R SH, Wiig JN,. Skin closure:-A prospective randomized study 1989 [Available from: https://pubmed.ncbi.nlm.nih.gov/2660322/.

23. Edwards DJ ER. Skin closure using nylon and polydioxanone: a comparison of results 1995 [

24. Cromi A GF, Gottardi A,. Cosmetic outcomes of various skin closure methods following cesarean delivery: a randomized trial 2010 [

25. Kundra RK NS, Saithna A,. Absorbable or non-absorbable sutures? A prospective, randomised evaluation of aesthetic outcomes in patients undergoing elective day-case hand and wrist surgery. 2010 [Available from: https://pubmed.ncbi.nlm.nih.gov/20659358/.

26. Osmond MH KT, Quinn JV,. Economic comparison of a tissue adhesive and suturing in the repair of pediatric facial lacerations. 1955 [Available from: https://pubmed.ncbi.nlm.nih.gov/7776090/.

27. Hargreaves J. Assessing the most clinically and cost effective method of closing skin following surgery. 2010 [Available from: https://pubmed.ncbi.nlm.nih.gov/20882833/.

28. Phillips JP EW. Laceration repair using rapid absorbing suture in the emergency department improves resource utilization: saving time and money. 2012

29. Cena Tejani ABS, Micheal D Rosen, Albert K Nakanishi, Robert G Flood, Mathew A Clott, Paul G Saccone, Raemma P Luck. A comparison of cosmetic outcomes of lacerations on the extremities and trunk using absorbable versus nonabsorbable sutures 2014 [Available from: https://pubmed.ncbi.nlm.nih.gov/25039547/.

30. A Bloemen PvD, B F Huizinga, A G M Hoofwijk,. Randomized clinical trial comparing polypropylene or polydioxanone for midline abdominal wall closure 2011 [Available from: https://pubmed.ncbi.nlm.nih.gov/21254041/.

31. Fernando Docobo-Durantez CS-P, Blas Flor-Civera, Salvador Lledó-Matoses, Esther Kreisler, Sebastiano Biondo,. Randomized clinical study of polydioxanone and nylon sutures for laparotomy clousure in high-risk patients 2006 [Available from: https://pubmed.ncbi.nlm.nih.gov/16753121/.

32. Mirza SM HF, Khalid K, Ali AA, Chaudry AM,. A prospective randomized trial of polypropylene and polydioxanone 2003 [

33. Murphy PG TE, Cross S,. Skin closure and the incidence of groin wound infection: a prospective study 1995 [Available from: https://pubmed.ncbi.nlm.nih.gov/8541198/.

34. L A Israelsson TJ. Closure of midline laparotomy incisions with polydioxanone and nylon: the importance of suture technique 1994 [Available from: https://pubmed.ncbi.nlm.nih.gov/7827883/.

35. A E Cameron CJP, E S Field, R C Gray, A P Wyatt. A randomised comparison of polydioxanone (PDS) and polypropylene (Prolene) for abdominal wound closure 1987 [Available from: https://pubmed.ncbi.nlm.nih.gov/3111339/.

36. Z H Krukowski ELC, J Engeset, N A Matheson,. Polydioxanone or polypropylene for closure of midline abdominal incisions: a prospective comparative clinical trial 1987 [Available from: https://pubmed.ncbi.nlm.nih.gov/3117165/.

37. J Wissing TJvV, M E Schattenkerk, H F Veen, R J Ponsen, J Jeekel,. Fascia closure after midline laparotomy: results of a randomized trial 1987 [Available from: https://pubmed.ncbi.nlm.nih.gov/3307992/.

38. D J Leaper AA, R E May, A P Corfield, R H Kennedy,. Abdominal wound closure: a controlled trial of polyamide (nylon) and polydioxanone suture (PDS) 1985 [Available from: https://pubmed.ncbi.nlm.nih.gov/3931536/.

